# Prevalence and risk factors for severe food insecurity and poor food consumption during a drought emergency in Ethiopia

**DOI:** 10.1101/2025.04.25.25326443

**Authors:** Noah Baker, Yunhee Kang, Gregory Makabila, Seifu Tadesse, Shannon Doocy

## Abstract

Frequent drought emergencies have heightened nutritional concerns in Ethiopia. This cross-sectional secondary data analysis assesses prevalence and risk factors of severe food insecurity and poor food consumption in Productive Safety Net Programme (PSNP) households in drought-prone Ethiopia. Data was from the USAID-funded Resilience Food Security Activity baseline survey. Severe food insecurity (n=4628; multivariate n=4335) was defined as Food Insecurity Experience Scale (≥7) and poor food consumption (n=4554; multivariate n=4268) was defined as Food Consumption Score (≤22). Logistic regression identified risk and protective factors from sociodemographic, economic, crops/livestock, and water/sanitation/hygiene variables. Severe food insecurity prevalence was 77.79% and poor food consumption prevalence was 69.74%. Risk factors for severe food insecurity included women/girls aged 15-19 years (Adjusted OR=1.79; 95% CI: 1.36-2.34), currently pregnant (1.52; 1.17-1.96), history of pregnancy (3.46; 2.76-4.33), cash-earning work (1.35; 1.12-1.61), daily-per-capita food consumption <1.61USD (2.98; 1.91-4.66), crop-planting (1.67; 1.31-2.13), and handwashing materials (3.83; 1.92-7.63); protective factors included raising livestock/crops (0.50; 0.42-0.60) and oxen (0.34; 0.26-0.45). Risk factors for poor food consumption included female household-head (1.44; 1.15-1.81) and household-head with no education (1.46; 1.18-1.79), daily-per-capita food consumption <1.61USD (4.00; 2.58-6.21), and financial service use (2.10; 1.69-2.59); protective factors included women aged 15-19 (0.59; 0.46-0.76) and 30-49 (0.76; 0.63-0.91), currently pregnant (0.57; 0.47-0.70), history of pregnancy (0.70; 0.55-0.89), crop-planting (0.57; 0.44-0.75), raising livestock/crops (0.40; 0.34-0.48) and oxen (0.68; 0.52-0.90). This study found vulnerable PSNP groups to be households with women that are/have previously been pregnant, no education, low economic status, and lack of livestock. PSNP should tailor education towards local climate-resistant crops and prioritize the livestock market, breeding, and survivability (specifically oxen), as crop production appears insufficient to maintain food security. The dual burden of food insecurity and low food consumption threatens current and future generations, and data-driven action can help progress towards the goal of zero hunger in Ethiopia.

## Introduction

Ethiopia has been historically susceptible to drought because of its location in the Horn of Africa, proximity to the equator, and proclivity to low rainfall.^1^ Climate change has severely impacted the Horn of Africa, with 4 major droughts over the past 15 years, the most recent being the longest recorded drought in decades (2020-2022). Higher drought frequency has led to five straight consecutive seasons of limited rainfall since 2020.^2^ As of early October 2022, the Horn of Africa was undergoing the worst drought in the past 70 years, with four rainy seasons of below average precipitation.^3^ The 2023 Global Humanitarian Overview estimated almost 20 million individuals were affected solely by climactic shocks in eastern and southern Ethiopia, with over 590,000 displaced.^4^ The Office for the Coordination of Humanitarian Affairs (OCHA) estimated that 10 million need food assistance and 3 million pregnant/lactating women (PLW) and children need nutritional support. Humanitarian assistance cannot cover these gaps, and in late 2022 the need for agriculture, food, health, and nutrition assistance respectively reached 52%, 82%, 12%, and 48% of the population in drought-affected areas.^5^

Historically, the East Hararghe Zone (EHZ) of Oromia has had high levels of household food insecurity.^6^ Insufficient rainfall and poor harvests in late 2022 led to emergency levels of food insecurity expected to extend through at minimum mid-2023.^7^ Famine Early Warning System (FEWS) data from November 2020-November 2022 detected that EHZ consistently experience “crisis” levels of food insecurity, and typically only drop to “stressed” with delivery of humanitarian aid.^8^ Many households are reliant on food distribution through humanitarian assistance or social protection schemes such as the Productive Safety Net Programme (PSNP). PSNP households depend upon this government assistance program that aims to support the poorest households in Ethiopia, which are at a higher risk for food insecurity.^9^ In EHZ, there were upwards of approximately 135,000 Internally Displaced People (IDP), primarily induced by conflict and climate.^10^ Displacement can heighten vulnerability to food insecurity and undernutrition, despite many displaced households targeted for food assistance. However, supply chain disruptions, attributed to wars in Tigray and between Russia and Ukraine, have restricted food assistance and contributed to food price volatility.^11^ The Protection Cluster in Ethiopia described high levels of severe food insecurity in EHZ attributed to drought conditions, floods, and pests, which have been compounded by recent conflict and the spread of cholera in four EHZ woredas.^12^ Humanitarian assistance has not been able to cover the nutritional gap and vulnerable populations such as women/girls of reproductive age (WRA) and children under 5 (CU5) are increasingly susceptible to negative nutritional outcomes.^8^

Although the Sustainable Development Goals (SDG) aim to reach zero hunger in Ethiopia by 2030, the most recent report indicates “major challenges” still remain to achieve this. Specifically, major challenges remain in reducing the magnitude of undernutrition, while child stunting has maintained a high prevalence.^13^ With increased drought frequency, high prevalence of food insecurity and low food consumption are likely to remain prevalent in EHZ. The persistent gap between food aid requirements and available assistance primarily experienced by PSNP households reinforces the need for research on PSNP households to develop tailored interventions and support progress toward the SDG of zero hunger. Several studies assess PSNP households, but these primarily evaluated PSNP program effectiveness on food security, food consumption, and asset accumulation, ^14 15 16 17^. Although these studies have demonstrated the positive impacts of PSNP, they maintain limited capacity to provide a large-scale analysis of rural PSNP households in drought emergencies, nor do they address food security and consumption in the context of religious fasting conditions and low availability seasons. There are also no studies that comparatively assess relationships between the consumption-based measure of Food Consumption Score and the perception-based measure of Food Insecurity Experience Scale (FIES). Much literature has assessed FCS as a measure of food security, however, the dimensions of food security consist of physical availability, access, utilization, and stability,^18^ which is difficult to quantify using consumption-based measures that may fluctuate daily. Therefore it is important to analyze this in combination with individual perceptions.^19^ The majority of contemporary food security and consumption research in Ethiopia focuses on general populations as opposed to PSNP households, who experience high poverty, lack resources, have limited access to services, and are among the most susceptible in drought emergencies. This analysis will highlight PSNP food insecurity and food consumption to support sustainable food security and resilience in drought-prone areas. Overall, this study aims to examine the prevalence of and risk factors for household severe food insecurity and poor food consumption in vulnerable PSNP households in Ethiopia during a drought emergency.^20^

## Materials and Methods

### Data Source

This secondary data analysis used data from a cross-sectional survey of PSNP households in eight of the 19 woredas in EHZ, Oromia, Ethiopia during a drought emergency. The survey was conducted as a baseline for the Ifaa project, a five-year USAID-funded Food Security and Resilience Activity (RFSA) implemented by a Catholic Relief Services-led consortium between 2022 and 2026. This study focuses on leveraging existing data to identify potential avenues to improve food security and food consumption in humanitarian emergency settings.

Ifaa aims to enhance food security and resilience and targets 4,683 PSNP households with 27,869 beneficiaries. The baseline survey was conducted May-June 2022 and assessed food insecurity and food consumption respectively in 4678 and 4601 households. The baseline survey included a sample of PSNP-eligible households in eight EHZ woredas (Babile, Chinaksen, Deder, Fedis, Gursum, Jarso, Melka Belo, and Midega Tola). Of the 241 eligible kebeles, 34 were excluded for security considerations and 11 kebeles because of selection for another study, leaving 196 kebeles. A stratified cluster design randomly selected 120 kebeles, and within each kebele, 39 PSNP households with a woman/girl of reproductive age (WRA, 15-49yrs) were randomly selected. This study’s sample assessed food insecurity and food consumption respectively in 4628 and 4554 households with descriptive statistics and univariate regression, and for multivariate regression respectively the samples were 4335 and 4268.^20^

The survey respondent was the WRA residing in the selected household, and in households with multiple, one was randomly selected. Details of the survey methodology are available in the Ifaa baseline survey report.^20^ WRA survey respondents provided information on household sociodemographics, economic activity, crop/livestock production, and WASH materials/practices. ^20^

### Outcomes

#### Severe Food Insecurity (SFI)

Using the FIES developed by the Food and Agriculture Organization (FAO), eight questions were asked to assess the difficulty of households in accessing adequate food.^21^ FIES is an indicator of food insecurity specifically tailored towards data collection in emergency nutrition settings and is commonly used by aid programs and the WFP.^22 23^ The nature of this measure aligns with this study’s objectives to assess food insecurity in drought emergency settings, and with the high caseload of SFI, the need for streamlined data collection tools such as FIES are needed to quickly and accurately assess population food security. Each question consisted of a binary indicator (1=Yes; 0=No), and these binary responses were summed to reach a raw score ranging from 0-8. Households were categorized by the FIES scores, with scores ranging from 4-6 indicating moderate food insecurity and 7-8 indicating severe food insecurity (SFI).^20^ Dummy variables were generated to assess factors associated with the binary outcome of severe food insecurity (SFI) when FIES ≥7. FIES is used in this study as a summary score to assess the aggregate effects of physical availability, access, utilization, and stability.

#### Poor Food Consumption

The household FCS developed by the WFP measures food group consumption frequency, dietary diversity, and nutritional importance^24^. Households were asked how many days a household consumed each of nine food groups in the past 7 days: The food groups included: Staples; Pulses; Vegetables; Fruit; Meat/Fish; Milk/Dairy; Sugar/Honey; Oil/Fats; and Condiments.^20^ Each food group was weighted by its nutritional value, and these scores were summed ranging from 0-112. Scores ranging from 0-21 indicate poor FCS, 22-35 borderline FCS, and 36-112 acceptable FCS. A binary variable of poor food consumption (FCS≤22) was generated for further analysis.

### Covariates

Covariates were selected through a literature review. Sociodemographic characteristics included (1) women’s age group (20-29; 15-19; 30-49), (2) number of children under 5 (CU5) in household (none; one; two or more), (3) currently pregnant vs. not pregnant, (4) history of pregnancy vs. no history of pregnancy, (5) household head’s no education vs. at least primary education, (6) WRA no education vs. at least primary education, and (7) married vs. not married. Household economic activity and crop/livestock production characteristics included (1) cash-earning work performed in the past year vs. not performed, (2) saved money vs. did not save (3) used financial services vs. did not use, (4) access to a land plot vs. no access, (5) raised livestock/cultivated crops with intent to sell vs. did not raise, (6) raised oxen/poultry/goats vs. did not raise, and (7) planted crops they make decisions over vs. did not plant). Water, Sanitation and Hygiene (WASH) materials/practices included (1) had handwashing materials vs. did not have, (2) correctly treated water vs. did not correctly treat, and (3) used improved sanitation facilities vs. did not use.

### Statistical Analysis

Exploratory data analysis was conducted to present n (%) of categorical variables for outcomes and risk factor variables. Inclusion criteria consisted of PSNP households with a WRA respondent 15-49 years of age. Logistic regression was used to identify risk factors significantly associated with the dependent variables (SFI and poor FCS). First, univariate logistic regression identified variables significant at the 95% confidence level. Variables significant at the 95% confidence level in univariate regressions were retained for the multivariate model. Covariates significant at the 95% confidence level (α=0.05) in multivariate models were considered risk/protective factors of SFI and poor FCS. Respondents with missing data were included in the descriptive and univariate regression analysis, but excluded from the multivariate regression analysis. Covariates that displayed collinearity were excluded from the analysis. Stata 17 (STATA Co.) was used for the statistical analysis.

### Ethical Approval

Causal Design attained Institutional Review Board (IRB) approval for Ifaa baseline data collection from the Ethiopian Society of Sociologists, Social Workers, and Anthropologists (ESSSWA).^20^ The study authors received an exempt determination from the Johns Hopkins Bloomberg School of Public Health IRB (FWA: #00000287).

## Results

The descriptive analysis was conducted using the SFI sample due to its greater sample size, as the maximum observed difference in variable proportions between SFI and poor FCS was minimal (0.43%), supporting the use of SFI for descriptive reporting. (Table 1; Table 2) Out of 4628 HH, the prevalence of severely food insecure households in this sample was 77.8% (Table 1), and out of 4554 HH, most (69.74%) reported poor food consumption (Table 2) (Figure 1). Out of 4628 households, 60.2% of HH were women aged 30-49 years old. 52.25% of households had one or more children under five. 13.4% of HH had pregnant women and 66.3% of women were married. 86% of mothers had no formal education and an almost equal percentage of HH heads (86.5%) also had no formal education. 28.8% of Women performed cash-earning work in the past 12 months. Nearly all HH (97.5%) spent <1.61 USD on daily-per-capita food consumption. A small proportion of households (18.4%) used financial services. 22.9% of households raised livestock/cultivated crops with intent to sell. The type of livestock varied for oxen (6.4%), poultry (19.6%), goats (23.5%), and planted crops they make decisions over (91.2%). Indicators related to WASH were overall low; only 2.4% had hand washing materials, 7.4% treated drinking water, and 15.9% used improved sanitation facilities (Table 1).

**Figure 1:**
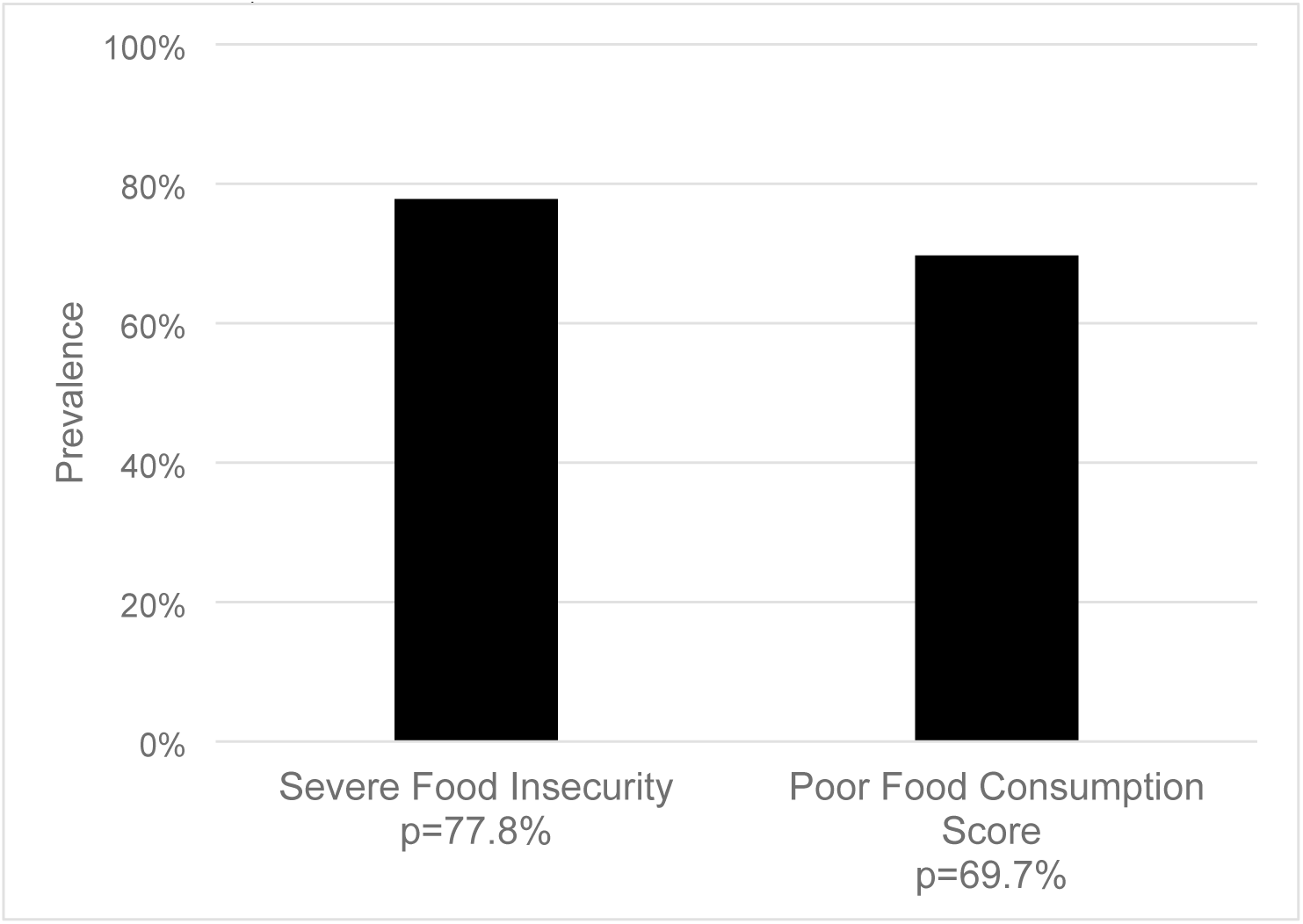
Prevalence of Severe Food Insecurity and Poor Food Consumption Score in East Hararghe Zone, Oromia, Ethiopia (Severe Food Insecurity n=4628; Poor Food Consumption Score n=4554)

**Figure 2:**
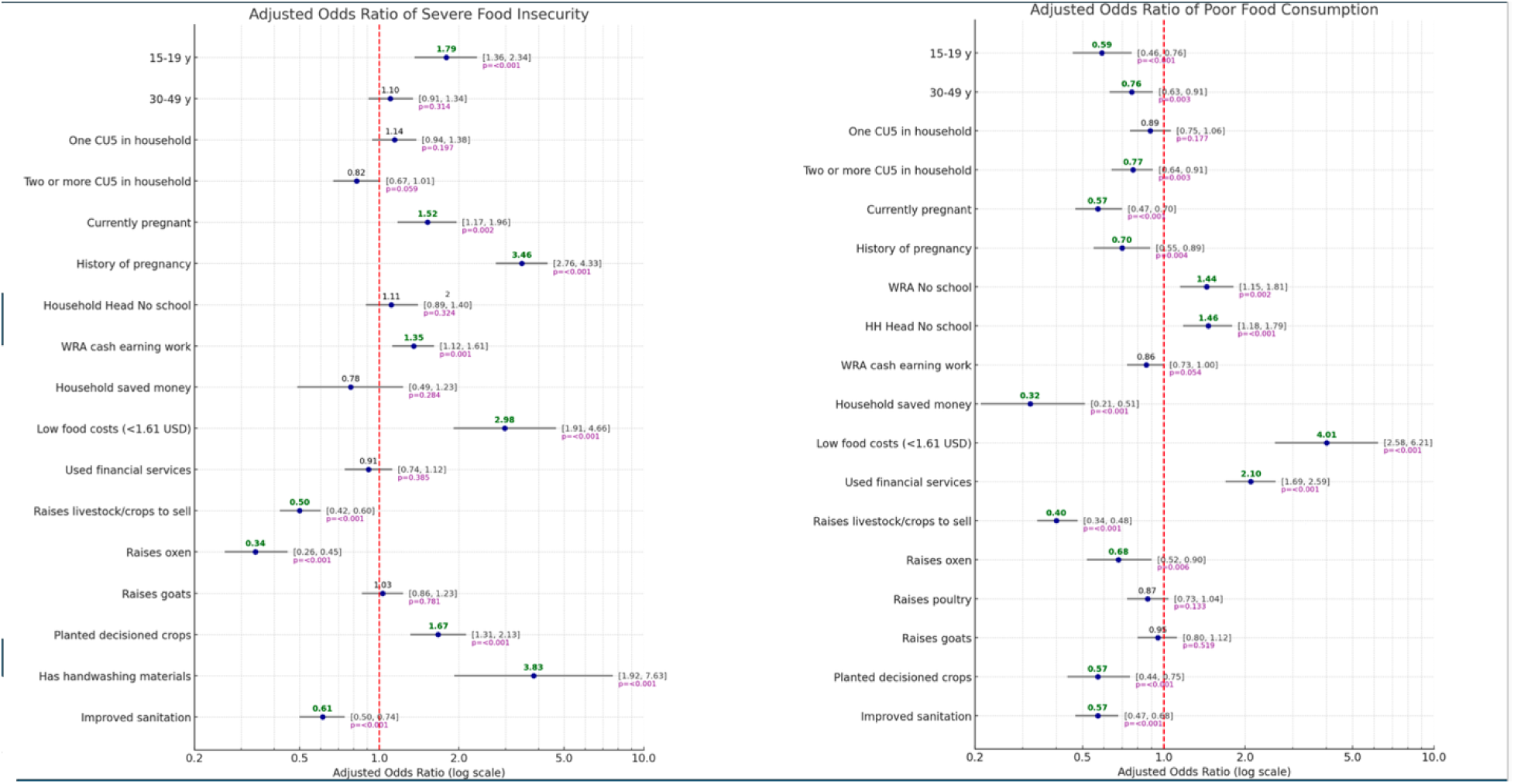
Results from multivariate logistic regression expressed in a Forest Plot. Factors associated with severe food insecurity and poor food consumption score at the 95% confidence level (α=0.05) (Severe Food Insecurity n=4335; Poor Food Consumption Score n=4268)

**Table 1.**
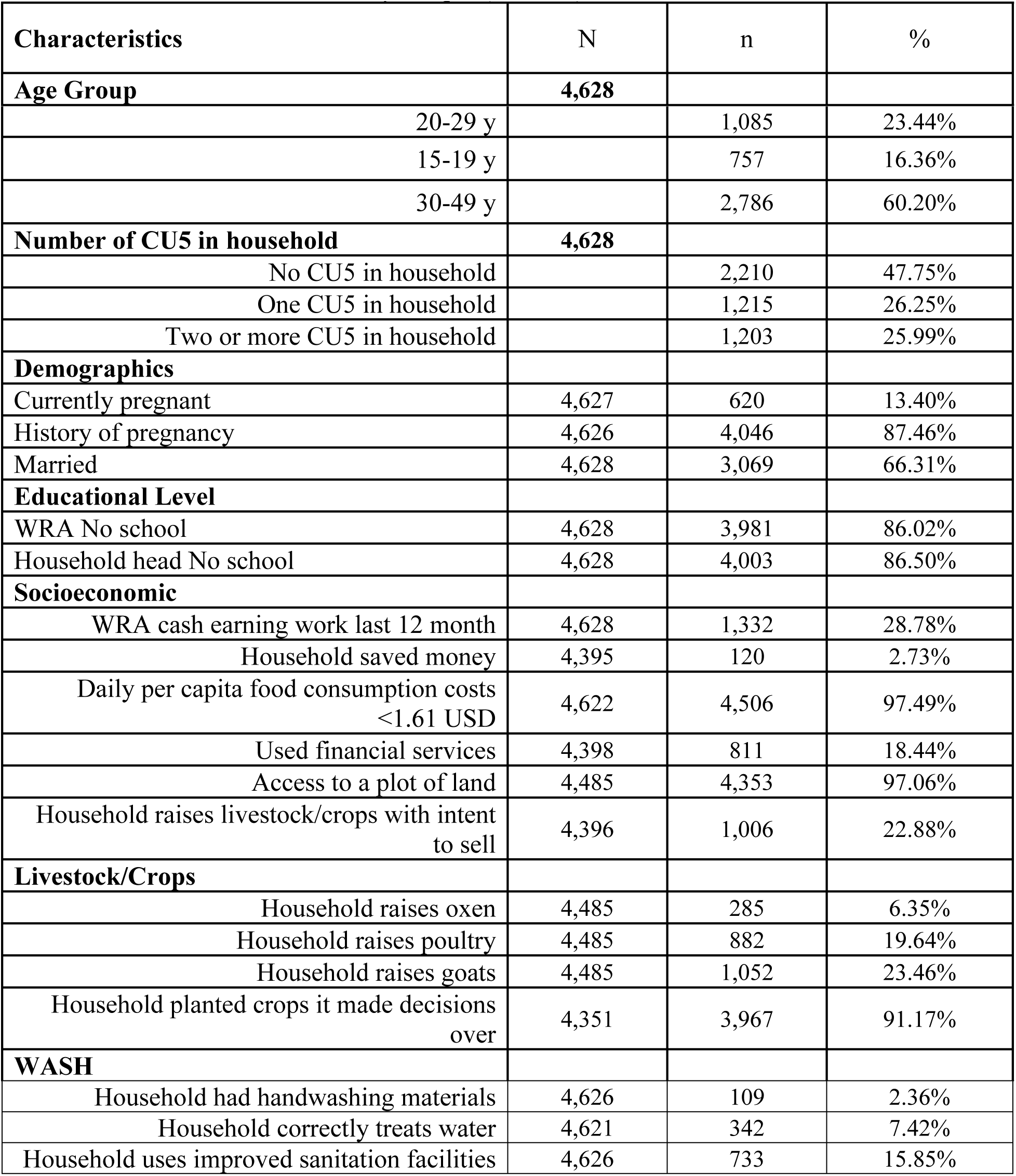
Characteristics of socio-demographics, economic activity, crop/livestock production, and WASH in Severe Food Insecurity sample (N=4628)

**Table 2.**
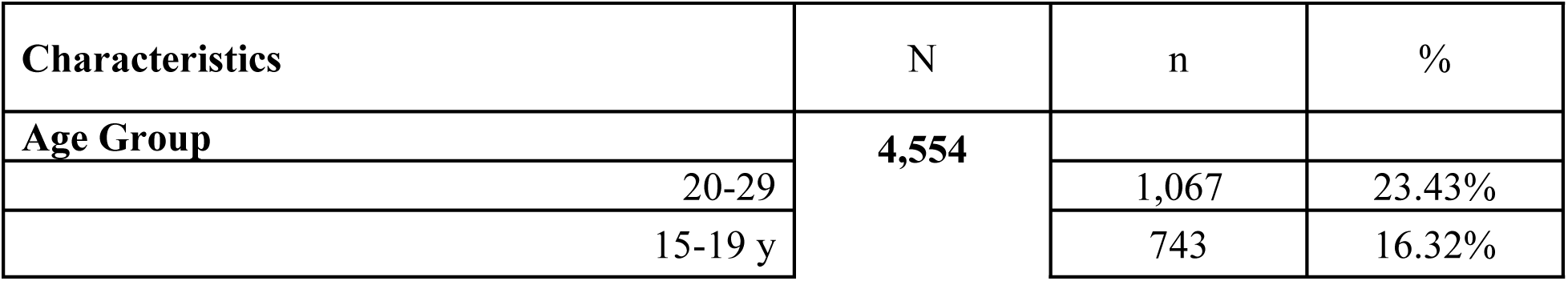
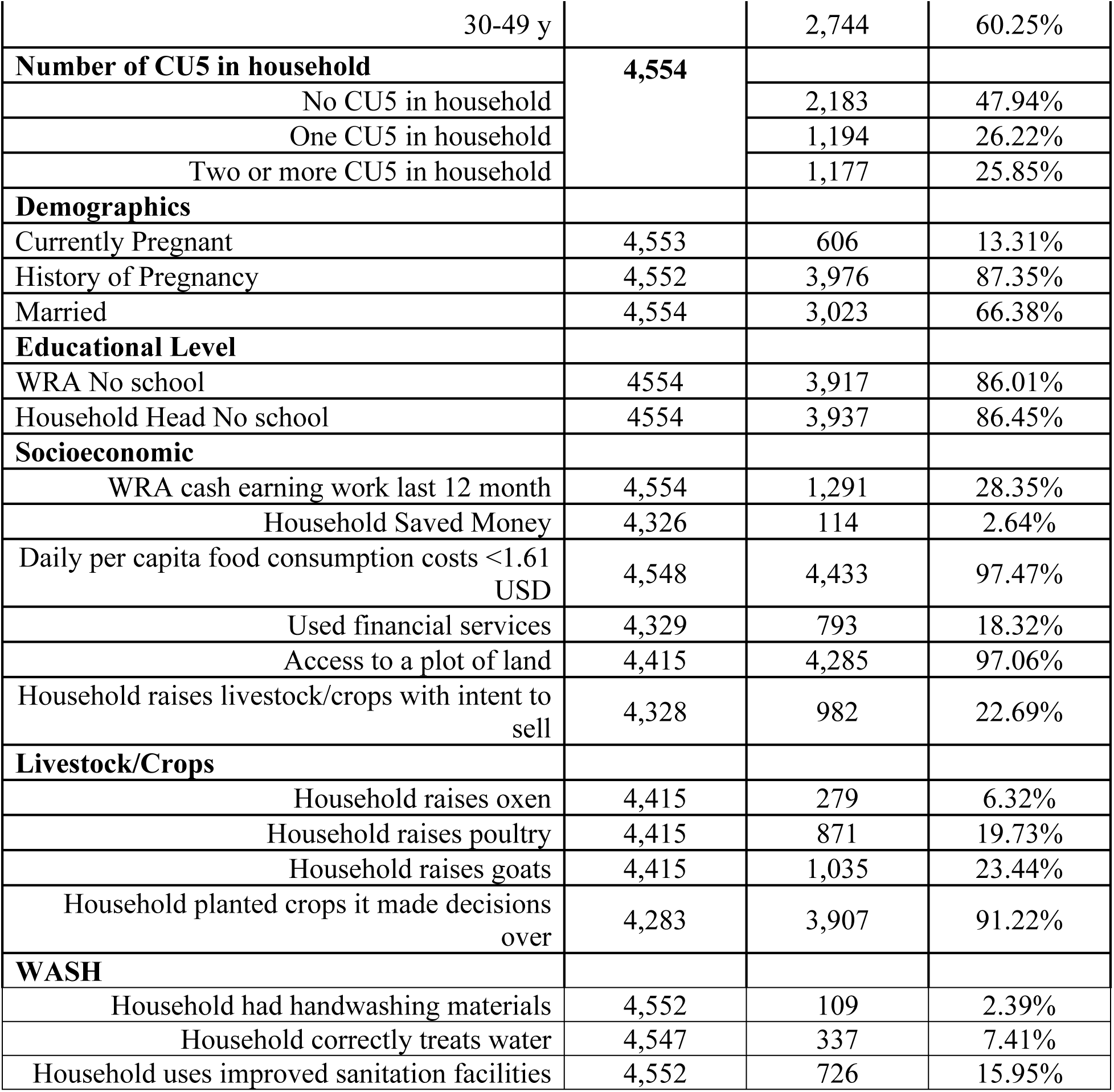
Characteristics of socio-demographics, economic activity, crop/livestock production, and WASH in Poor Food Consumption sample (N=4554)

| Characteristics | N | n | % |
| --- | --- | --- | --- |
| **Age Group** | **4,554** |  |  |
| 20-29 |  | 1,067 | 23.43% |
| 15-19 y |  | 743 | 16.32% |
| 30-49 y |  | 2,744 | 60.25% |
| **Number of CU5 in household** | **4,554** |  |  |
| No CU5 in household |  | 2,183 | 47.94% |
| One CU5 in household |  | 1,194 | 26.22% |
| Two or more CU5 in household |  | 1,177 | 25.85% |
| **Demographics** |  |  |  |
| Currently Pregnant | 4,553 | 606 | 13.31% |
| History of Pregnancy | 4,552 | 3,976 | 87.35% |
| Married | 4,554 | 3,023 | 66.38% |
| **Educational Level** |  |  |  |
| WRA No school | 4554 | 3,917 | 86.01% |
| Household Head No school | 4554 | 3,937 | 86.45% |
| **Socioeconomic** |  |  |  |
| WRA cash earning work last 12 month | 4,554 | 1,291 | 28.35% |
| Household Saved Money | 4,326 | 114 | 2.64% |
| Daily per capita food consumption costs <1.61 USD | 4,548 | 4,433 | 97.47% |
| Used financial services | 4,329 | 793 | 18.32% |
| Access to a plot of land | 4,415 | 4,285 | 97.06% |
| Household raises livestock/crops with intent to sell | 4,328 | 982 | 22.69% |
| **Livestock/Crops** |  |  |  |
| Household raises oxen | 4,415 | 279 | 6.32% |
| Household raises poultry | 4,415 | 871 | 19.73% |
| Household raises goats | 4,415 | 1,035 | 23.44% |
| Household planted crops it made decisions over | 4,283 | 3,907 | 91.22% |
| **WASH** |  |  |  |
| Household had handwashing materials | 4,552 | 109 | 2.39% |
| Household correctly treats water | 4,547 | 337 | 7.41% |
| Household uses improved sanitation facilities | 4,552 | 726 | 15.95% |

### Severe Food Insecurity

Households with WRA aged 15-19 years were 1.79 times more likely to experience SFI (95% CI: 1.36, 2.34) when compared to WRA 20-29 years of age. Households with the WRA respondent pregnant were 1.52 times more likely to experience SFI than households without pregnant women (95% CI: 1.17, 1.96). Households with WRA with a history of ever being pregnant had a 3.46 times higher likelihood of SFI (95% CI: 2.76, 4.33). Performing cash-earning work throughout the previous year was more likely and maintaining daily per capita food expenditure <1.61 USD were associated with experiencing SFI (OR=1.35; 95% CI: 1.12, 1.61 and OR=2.98; 95% CI: 1.91, 4.66, respectively). Raising oxen (OR=0.34; 95% CI: 0.26, 0.45) or raising livestock or cultivated crops (OR=0.50; 95% CI: 0.42, 0.60) were less likely to experience SFI. Households that planted crops were 1.67 times more likely to experience SFI (95% CI: 1.31, 2.13). However, 92.48% of severely food insecure households planted crops, while 34.98% raised target livestock (oxen, poultry, goats) indicating these results were skewed from the proportion of households that planted crops and also raised livestock. Households that retained handwashing materials were 3.83 times more likely to experience SFI (95% CI: 1.92, 7.63). Households that utilize improved sanitation facilities were 0.61 times less likely to experience SFI (95% CI: 0.50, 0.74) (Table 3).

**Table 3.** Factors associated with Severe Food Insecurity from univariate (n=4628) and multivariate logistic regression (n=4335) among PSNP households in East Hararghe Zone, Oromia, Ethiopia. Crude and Adjusted odds ratios displayed.

| Characteristics | n | Univariate OR | 95%CI | p | Multivariate OR (n=4335) | 95%CI | p |
| --- | --- | --- | --- | --- | --- | --- | --- |
| **Age Group** | 4,628 |  |  |  |  |  |  |
| 20-29 y (Ref) |  | 1.00 |  |  | 1.00 | . | . |
| 15-19 y |  | 1.33 | (1.06, 1.66) | 0.012 | 1.79 | (1.36, 2.34) | <0.001 |
| 30-49 y |  | 1.29 | (1.10, 1.52) | 0.002 | 1.10 | (0.91, 1.34) | 0.314 |
| **Number of CU5 in household** | 4,628 |  |  |  |  |  |  |
| No CU5 in household (Ref) |  | 1.00 |  |  | 1.00 | . | . |
| One CU5 in household |  | 1.03 | (0.87, 1.22) | 0.749 | 1.14 | (0.94, 1.38) | 0.197 |
| Two or more CU5 in household |  | 0.77 | (0.64, 0.94) | 0.008 | 0.82 | (0.67, 1.01) | 0.059 |
| **Demographics** |  |  |  |  |  |  |  |
| Currently pregnant (Ref: No) | 4,627 | 1.72 | (1.37, 2.17) | <0.001 | 1.52 | (1.17, 1.96) | 0.002 |
| History of pregnancy (Ref: No) | 4,626 | 2.68 | (2.23, 3.22) | <0.001 | 3.46 | (2.76, 4.33) | <0.001 |
| Married (Ref: No) | 4,628 | 0.91 | (0.78, 1.05) | 0.196 | . | . | . |
| **Educational Level** |  |  |  |  |  |  |  |
| WRA No school (Ref: formal education) | 4,628 | 0.99 | (0.81, 1.21) | 0.942 | . | . | . |
| Household Head No school (Ref: formal education) | 4,628 | 1.29 | (1.07, 1.57) | 0.009 | 1.12 | (0.89, 1.40) | 0.324 |
| **Socioeconomic** |  |  |  |  |  |  |  |
| WRA cash earning work last 12 months (Ref: No) | 4,628 | 1.4 | (1.19, 1.64) | <0.001 | 1.35 | (1.12, 1.61) | 0.001 |
| Household saved money (Ref: No) | 4,395 | 0.51 | (0.35, 0.75) | 0.001 | 0.78 | (0.49, 1.23) | 0.284 |
| Daily per capita food consumption costs <1.61 USD (Ref: >=1.61USD?) | 4,622 | 2.73 | (1.88, 3.97) | <0.001 | 2.98 | (1.91, 4.66) | <0.001 |
| Used financial services (Ref: No) | 4,398 | 0.67 | (0.56, 0.80) | <0.001 | 0.91 | (0.74, 1.12) | 0.385 |
| Access to a plot of land (Ref: No) | 4,485 | 1.13 | (0.75, 1.70) | 0.551 | . | . | . |
| Household raises livestock/crops with intent to sell (Ref: No) | 4,396 | 0.49 | (0.42, 0.58) | <0.001 | 0.50 | (0.42, 0.60) | <0.001 |
| **Livestock/Crops** |  |  |  |  |  |  |  |
| Household raises oxen (Ref: No) | 4,485 | 0.33 | (0.26, 0.43) | <0.001 | 0.34 | (0.26, 0.45) | <0.001 |
| Household raises poultry (Ref: No) | 4,485 | 1.07 | (0.90, 1.28) | 0.464 | . | . | . |
| Household raises goats (Ref: No) | 4,485 | 0.85 | (0.72, 1.00) | 0.048 | 1.03 | (0.86, 1.23) | 0.781 |
| Household planted crops it made decisions over (Ref: No) | 4,351 | 1.91 | (1.52, 2.39) | <0.001 | 1.67 | (1.31, 2.13) | <0.001 |
| **WASH** |  |  |  |  |  |  |  |
| Household has handwashing materials (Ref: No) | 4,626 | 2.59 | (1.38, 4.85) | 0.003 | 3.83 | (1.92, 7.63) | <0.001 |
| Household correctly treats water (Ref: No) | 4,621 | 1.14 | (0.87, 1.50) | 0.344 | . | . | . |
| Household uses improved sanitation facilities (Ref: No) | 4,626 | 0.63 | (0.52, 0.75) | <0.001 | 0.61 | (0.50, 0.74) | <0.001 |

### Poor Food Consumption Score

When compared to WRA 20-29 years, households of WRA aged 15-19 or WRA 30-49 years were less likely to reside in households with poor food consumption (OR=0.59; 0.46, 0.76 and OR=0.76; 0.63, 0.91, respectively). Households with heads without formal education were 1.46 times more likely (1.18, 1.79) to have poor food consumption, and households of WRA without formal education were 1.44 times more likely to have poor food consumption (1.15, 1.81) compared to WRA with higher education. Households with currently pregnant WRA or WRA with a history of pregnancy were less likely to experience poor food consumption (OR=0.57; 0.47, 0.70; OR=0.70; 0.55, 0.89, respectively). Households that planted crops they made decisions over were less likely to consume poor food consumption (OR=0.57; 0.44, 0.75). Households that saved money were less likely to consume poor food consumption (OR=0.32; 0.21, 0.51). Households that used financial services were 2.10 times more likely to have poor food consumption (1.69, 2.59). Households with per capita food consumption <1.61 USD were 4.01 times more likely to have poor food consumption (2.58, 6.21). Households that raised oxen were less likely to have poor food consumption (OR=0.68; 0.52, 0.90) compared to households not raising oxen. Cultivating crops/livestock with intent to sell were negatively associated with poor food consumption (OR=0.40; 0.34, 0.48). Households that used improved sanitation facilities were less likely to have poor food consumption (OR=0.57; 0.47, 0.68) (Table 4).

**Table 4:**
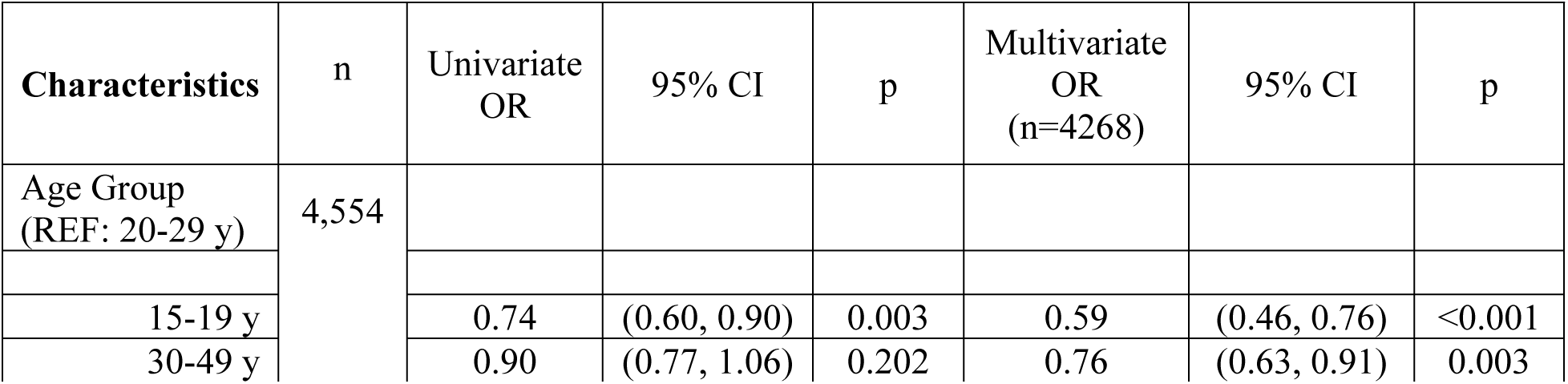
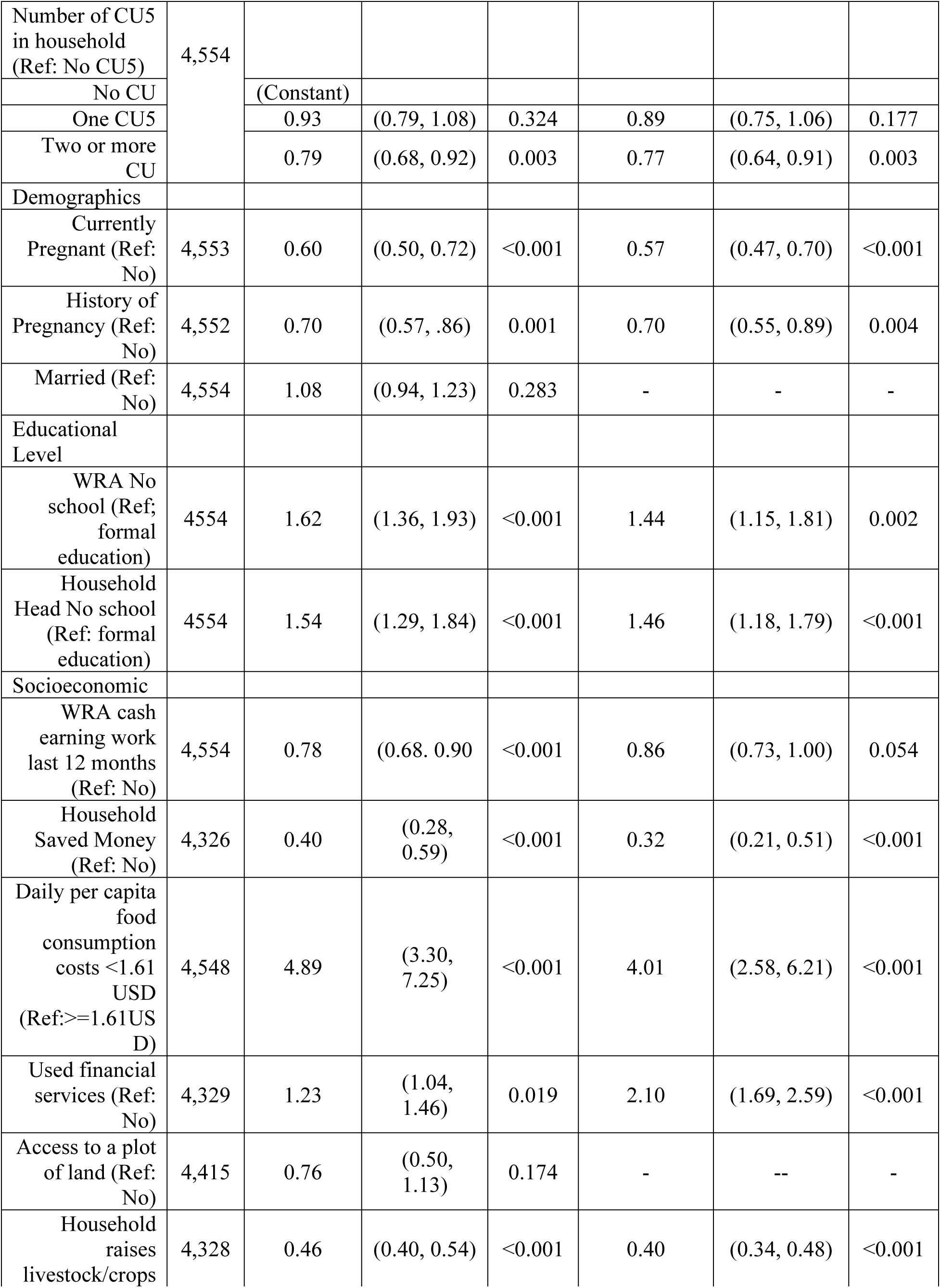
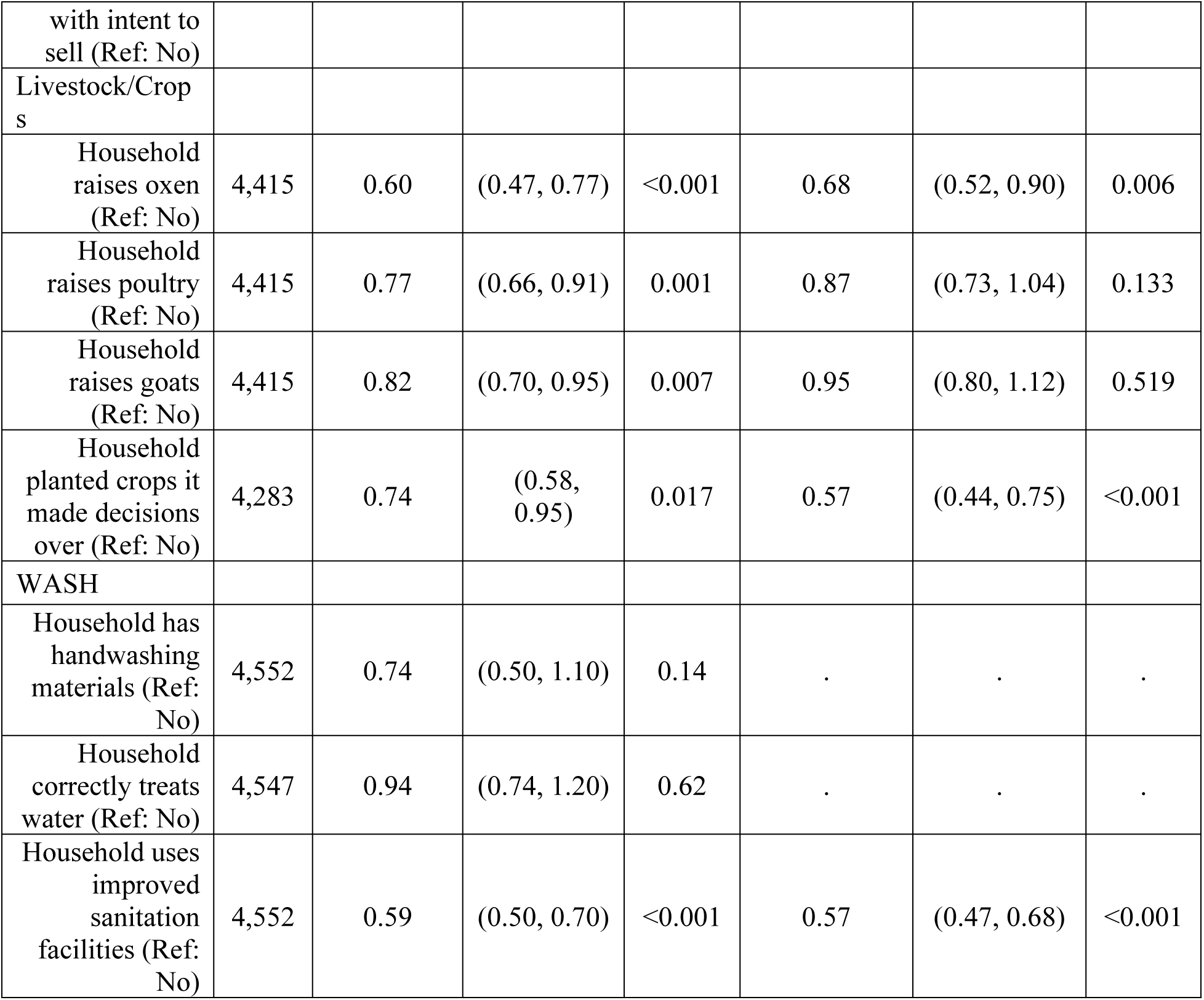
Factors associated with Poor Food Consumption Score from univariate (n=4554) and multivariate logistic regression (n=4268) among PSNP households in East Hararghe Zone, Oromia, Ethiopia. Crude and Adjusted odds ratios displayed

## Discussion

Underlying the importance of this study, most relevant research assessed the general population, while this study specifically assesses populations with known high food insecurity and low food consumption to determine actionable inferences that decrease the risk of nutritional morbidity related to persistent exposure to emergency drought conditions. Accordingly, this study revealed a high prevalence of severe food insecurity (77.8%) and poor food consumption (69.7%) among Productive Safety Net Programme (PSNP) households in East Haraghe Zone. The risk factors for severe food insecurity included households with WRA aged 15-19, currently pregnant, history of pregnancy, cash-earning work, daily-per-capita food consumption < 1.61 USD, crop-planting, and owning handwashing materials. Protective factors for severe food insecurity included households with WRA raising livestock/crops with the intent to sell, raising oxen, and using improved sanitation facilities. Risk factors of poor food consumption included households with WRA without formal education, household head without formal education, daily-per-capita food consumption < 1.61 USD, and financial service use. The protective factors for poor food consumption were households with WRA aged 15-19, two or more CU5 in the household, currently pregnant, history of pregnancy, households saving money, raising livestock/crops with intent to sell, and raising oxen.

The prevalence of food insecurity in Fedis woreda, EHZ was estimated to be 58% by Mulugeta et al, ^25^ and Getaneh et al identified food insecurity prevalence in Rift Valley as 68%. ^26^ A study by Aweke et al of EHZ smallholder farms identified the prevalence of poor food consumption in the preharvest season as 21.9%^27^, while Fite et al in Haramaya detected unacceptable food consumption to be 45.5%.^28^ These studies were conducted before intensified drought and record-breaking consecutive seasons of low rainfall ^2^, which likely contributed to the higher magnitude of severe food security and poor food consumption found in this sample. Additionally, a study by Hiruy et al in southwest Oromia collected data from November to December 2021, deeper into the rainy season that still displayed levels of food insecurity at 62.4%, with 28.1% of the sample experiencing severe food insecurity.^29^ Another study by Markos et al in southwest Oromia noted 16.4% poor FCS^30^, while a study of poor households by Dula in Gelan City identified poor FCS to be 58.33%.^31^ This emphasizes the importance of focusing academic research on poor households in drought emergency settings, particularly in highly affected regions such as East Oromia and Somali, as food insecurity and poor food consumption vary depending on the context.

### Sociodemographic Characteristics

EHZ is primarily rural, and since women with a history of pregnancy or currently pregnant were risk factors for SFI, this aligns with the Areba et al study that detected pregnant women from rural areas were more likely to be food insecure.^32^ Interestingly, although women with a history of pregnancy or currently pregnant in this sample were risk factors for SFI, these factors exhibited protective associations with poor FCS. FIES provides a perceptual measure of food security, while the consumption-based measure of FCS provides more insight on nutrient intake and dietary diversity. It is important to assess concordance between these two measures, as consumption-based measures may fluctuate daily in food-insecure individuals, and perception of food security status is based on continual experience. This could be attributed to a range of factors, such as community support or programming that helps PLW obtain adequate consumption. However, this may not be reflected in overall food security status. This indicates factors that alter perception of food security such as pregnancy and children, intrahousehold food distribution, and community support for PLW/mothers in food-insecure households should be further explored. Male-headed households could also be a factor, as males may participate in income-generating activities and farming that could increase food consumption for their pregnant partners.^29^ As pregnant women were at a higher risk for SFI and lower risk for poor FCS, future studies on PLW should prioritize collecting MUAC in tandem with food insecurity and consumption data to determine influential relationships between the perceptions, consumption, and validated nutritional measures.

Similarly, WRA aged 15-19 years was a risk factor for SFI, but a protective factor for poor FCS. The detection of WRA aged 15-19 as a risk factor for SFI is comparable to a Jebena et al study that found an increased likelihood of food insecurity in female adolescents compared to males.^33^ Contrarily, in this sample, WRA aged 15-19 and 30-49 were protective factors for poor FCS compared to WRA aged 20-29. WRA with two or more CU5 in the household were protective factors for poor FCS. Household heads and WRA with no formal education were risk factors for poor FCS, and this aligns with Sisay et al where household heads with education were positively associated with acceptable FCS.^34^ In the Hiruy et al study, women with formal education increased the likelihood of food security, which conflicts with this study but aligns with the results in the food consumption sample.^29^ This indicates that increased education may heighten awareness of individual food consumption needs, but awareness alone is likely insufficient to increase the food security status of households. Since 86.01% of WRA and 86.45% of household heads in the poor FCS sample did not receive any schooling, this highlights concerns for education in EHZ.

### Economic activity

Performing cash-earning work in the past year was a risk factor for SFI, indicating income-generating activities may be insufficient to enable food security in drought emergencies. This conflicts with Areba et al, where employed pregnant women were less likely to be food insecure ^32^, and Getacher et al, where lactating mothers who did not participate in income-generating activities were more likely to be food insecure.^35^ While households that saved cash were protective factors for poor FCS, cash-earning work was a risk factor for SFI. This may indicate that cash-earning work was insufficient to maintain food security and that households with the capacity to save cash may allocate it towards food for consumption. But this may not affect the food security status of the household since saving cash may be unsustainable, and available funds could be allocated towards food for consumption during shocks/stressors. This potentially explains the positive association between cash-earning work and SFI, and the negative (albeit not statistically significant; p=0.054) association between cash-earning work and poor FCS. It should be noted that FIES is a measure based on individual perception, while FCS is a quantified measure of food consumption, diversity, and frequency, and is weighted by nutrient value. Therefore, food security status could potentially differ based on WRA perception, but with relatively similar prevalence, perceptions of household food security likely align with poor FCS. This inference is furthered since 82.76% of households with poor FCS were also severely food insecure, an important finding for surveillance purposes. Since daily-per-capita food consumption costs ≤1.61 USD was a risk factor for both SFI and poor FCS, and using financial services was a risk factor for poor FCS, households may not have the financial capacity to repay credit while maintaining food security and adequate food consumption. Since this conflicts with a study by Wubetie et al in southern Ethiopia, it complicates the impact of financial services such as credit on daily food consumption.^36^ But this furthers the inference that in EHZ, limited financial resources are a primary factor that affects food security and consumption. Since opportunities to increase financial resources are limited in drought-prone environments, humanitarian programming must fill the gap.

### Livestock/Crop production

Households that raised livestock and/or crops with the intent to sell and those that raised oxen both displayed distinctly protective associations with SFI and poor FCS. The protective relationship of raising livestock/crops with the intent to sell is similar to results from the Getacher et al study.^35^ aboveHouseholds that raised livestock/crops with the intent to sell was a protective factor for SFI, but importantly, this association is skewed by households that own livestock since 91.2% of households plant crops (Table 1). Specifically, oxen were protective factors for severe food insecurity. Since oxen were only raised by 6.35% of the SFI sample and 6.32% of the poor FCS sample, this likely indicates oxen are inaccessible to many households with insufficient financial capacity. This is consistent with the assessment by Gebissa et al,^37^ and the Mulugeta et al study also conducted in EHZ that identified oxen as facilitators of food security.^25^ While raising livestock, specifically oxen, saw protective associations, households that planted crops they made decisions over were more likely to be severely food insecure. This is inconsistent with the Getacher study where home gardens reduce the risk of food security,^35^ however, aligns with the Gebissa study where livestock ownership was positively associated with food security, while land cultivation saw no relationship.^37^ However, households that planted crops they made decisions over were a risk factor for SFI and a protective factor for poor FCS. This indicates planting crops may improve FCS by increasing availability and diversity of food to supplement diets, but may be insufficient to improve food security at the household level.

While the Aweke study found growing crops was a protective factor for poor FCS, increased livestock and farm income was also a protective factor, similar to the protective association in this sample between livestock/crop cultivation with intent to sell and planting crops the household made decisions over.^27^ Fite et al in Haramaya found women that who did not own agricultural land plots were negatively associated with acceptable FCS. ^28^Although there was no association with land plot access in this sample, these results are similar to the protective association of planting crops. ^28^ Important staple crops such as tef, maize, and sorghum all decreased during drought conditions in a study by Temam et al., on the implications of drought in dryland Ethiopia.^38^ Agricultural production in Oromia has diminished from drought conditions^39^, and since planting crops was a risk factor for severe household food insecurity, crop production does not appear to be sufficient to maintain food security. Livestock ownership is likely a better facilitator of food security, an inference also ascertained by Getaneh.^26^ Sileshi et al found rural households in EHZ with an increased number of livestock owned were negatively associated with poor/borderline FCS, which aligns with the protective relationship of raising livestock/cultivating crops with the intent to sell and raising oxen with poor FCS in this sample.^40^ This also aligns with the study by Sisay et al that detected livestock were positively associated with acceptable FCS.^34^ A study in northern Ethiopia by Vaitla identified that increased livestock units was positively associated with FCS, similar to the decreased likelihood of poor FCS when the household raised oxen and raised livestock/cultivated crops with the intent to sell.^41^ The results highlight that livestock, specifically oxen, are key facilitators of food security and that crop production can be a tool to supplement household diet. The protective and risk factors detected must be accounted for in the design of food assistance and nutrition programs.

However, livestock are more expensive to obtain and maintain than crops, and this investment is a higher risk in drought settings since livestock death could diminish household resilience.^5^ Livestock interventions could benefit from the climate-sensitive approach to agriculture to decrease the risk of death. Interventions to improve survivability and maintain the health of livestock could be another approach to improve livelihoods and economic capacity to facilitate food security and mitigate shocks/stressors. This is furthered by Tofu et al that found pastoralists had lower resilience than agro-pastoralists.^42^ The protective association displayed by households that raised oxen in this sample and contemporary literature indicates that scaling up oxen breeding may heighten availability, enhance the livestock market, and improve food security and resilience. It has been determined by Getahun et al that with the agriculture uncertainty in Ethiopia, income diversification for farming households through livestock, forest, or non-farm means may be beneficial.^43^ The relationship between sources of income diversification such as forest farming, and cash crops and its impact on food security should be further explored.

### Water, Sanitation and Hygiene

Households that used improved sanitation were protective factors for SFI and poor FCS. Contrary to this sample, Vaitla et al did not find improved sanitation to be associated with FCS.^41^

### Recommendations

Based on our findings, we recommend directing PSNP resources towards households with pregnant/lactating women, mothers, young women, no education, low economic status, and lack of livestock ownership. It is also recommended to leverage the ease of collecting food security and food consumption data for pregnant/lactating women in households that need food aid to direct resources toward improving both perceptual and consumption-based measures.^32^ The policymakers and implementing agencies could consider creating paid PSNP programs that leverage the knowledge of experienced farmers to geographically tailor education and enhance awareness of locally available nutrient-dense and climate-resistant crops.^34^ We recommend prioritizing livestock survivability and developing targeted programs that focus on increasing the population of livestock through breeding to improve the livestock market and create expanded opportunities.^25 26 27 36 37 40 41^ It can be a useful approach to applying PSNP behavioral change to reinforce the allocation of financial resources towards protein-rich foods and utilize farming to diversify diets with fruits/vegetables/nuts and generate supplementary income. ^43^ Lastly, we advise integrating food security programming and outreach with appropriate WASH practices to diminish communicable disease burden and diarrheal infections that are deleterious to nutritional status.

## Strengths and Limitations

As this is a secondary data analysis of data collected during the COVID-19 pandemic, there were no anthropometric measurements taken and this study is limited to assessing food security and food consumption. Middle Upper Arm Circumference (MUAC) is commonly used to assess nutrition in PLW, however this study broadly assesses food security and consumption in households with WRA. Other food security measures such as HFIAS may have provided more information on specific aspects of food security, but in an emergency setting, FIES is a more efficient measure. This is a cross-sectional study and cannot assess causality. Data collection occurred during fasting periods, and although this likely impacted WRA food consumption, it provides important data during times of fasting. The sample consists of PSNP beneficiaries, and low-income populations in Ethiopia that receive governmental support, which complicates inferences to the general population but enriches current literature on how to aid those most in need. Data was collected by personal recall and may be susceptible to recall bias. A common issue with studies that employ personal recall is the potential for social desirability bias exhibited by participants. While seasonality is not assessed in this study, food security and food consumption must be analyzed during periods of low consumption. Although a large sample size increases the likelihood of significant associations, this is also an indicator of appropriate covariate selection. It is important to note that sample sizes slightly differ by respectively a 6.54% and 6.48% difference between SFI and poor FCS univariate and multivariate models.

## Conclusion

The high prevalence of severe food insecurity (77.79%) and poor food consumption (69.74%) indicates a sizeable number of households in EHZ may be nutritionally susceptible. Importantly, this study identified vulnerable PSNP groups such as households with pregnant/lactating women, mothers, young women, no education, low economic status, and lack of livestock ownership. Notably, households with currently pregnant women or those with a history of pregnancy were more likely to experience SFI, but less likely to experience poor FCS. This relationship highlights a critical distinction that must be reflected in literature, food security is a perceptual, experience-based measure, while food consumption is a quantitative metric subject to daily variation based on community and environmental factors. Therefore, it is critical that both data points are collected to better contextualize evidence. The variation in factors associated with food insecurity and consumption suggest individual definitions of household food security may change depending on the presence of PLW, children, and household size. The relative feasibility of collecting FIES food insecurity data in settings where anthropometric measurement is impractical may be an effective surveillance tool to detect pregnant/lactating women and households that need food aid. With zonal schooling concerns, programs that leverage experienced farmers to geographically tailor education can enhance awareness of local climate-resistant crops to prioritize. Households that raised oxen and raised livestock/cultivated crops with the intent to sell displayed protective associations, and PSNP should prioritize the livestock market, breeding, and survivability (specifically oxen). Although crop production appears insufficient to maintain food security, this may facilitate adequate food consumption. With limited food availability, it may be beneficial to direct behavioral change efforts to allocate financial resources towards protein-rich foods, while employing farming to diversify diets with fruits/vegetables/nuts and generate supplementary income. Expanding livelihood pathways with innovative methods to diversify income may also support food security in the zone. The low utilization of WASH materials in EHZ threatens child health and nutrition, and integrating food security programming with appropriate WASH practices for agro-pastoralists may be beneficial. The dual burden of severe food insecurity and poor food consumption poses a threat to current and future generations in EHZ, and it is critical to take data-driven action to progress toward the sustainable development goal of zero hunger in Ethiopia.

## CONTRIBUTIONS

N.B. reviewed literature, designed study, analyzed data, interpreted results, drafted and finalized manuscript; Y.K. guided statistical analysis, assisted study design, reviewed and edited manuscript; GM provided dataset, reviewed the manuscript and provided feedback; ST reviewed manuscript and provided feedback; and SD oversaw study design, reviewed analysis, and reviewed and edited manuscript. Casual Design collected data.

## DATA AVAILABILITY

The dataset used in this study is not publicly available, as it is subject to third-party ownership by Catholic Relief Services. Access may be shared upon reasonable request, pending approval from the authors and Catholic Relief Services. Access to analytic code may also be shared upon reasonable request. The dataset for this study was the baseline survey of the USAID-funded Ifaa (RFSA) Resilience Food Security Activity implemented by a consortium led by Catholic Relief Services. (“Causal Design, IMPEL. Baseline study of the Ifaa Resilience Food Security Activity in Ethiopia. Vol. 1. Washington, DC: The Implementer-Led Evaluation & Learning Associate Award; 2022.”) Further information regarding the dataset is included in the *Materials and Methods* section. With any questions please reach out to the corresponding author Yunhee Kang or Noah Baker.

---

**Supplemental Table 1.** Indicator Explanation.

| **Covariates** | **Explanation** |
| --- | --- |
| **Age Group** | Used to assess difference in statistical relationship between age groups of WRA in households and outcomes |
| Female 20-29: Reference Group |  |
| Female 15-19 |  |
| Female 30-49 |  |
| **Number of CU5 in household** | Used to assess difference in statistical relationship between number of children in household and outcomes |
| No CU5 in household: Reference Group |  |
| One CU5 in household |  |
| Two or more CU5 in household |  |
| **Demographics** |  |
| Currently Pregnant | Used to assess statistical relationship between households with WRA that is currently pregnant and outcomes |
| History of Pregnancy | Used to assess statistical relationship between households with WRA history of being pregnant and outcomes |
| Marital Status: Married | Used to assess statistical relationship between households with married WRA and outcomes |
| **Educational Level** |  |
| WRA No school | Used to assess statistical relationship between households with WRA that did not receive education and outcomes |
| Household Head No school | Used to assess statistical relationship between household heads with that did not receive education and outcomes |
| **Socioeconomic** |  |
| Cash earning work last 12 months | Used to assess statistical relationship between households that performed cash-earning work in the last 12 months and outcomes |
| Household Saved Money | Used to assess statistical relationship between households that saved money in the last 12 months and outcomes |
| Daily per capita food consumption costs <1.61 USD | Used to assess statistical relationship between households with food consumption costs less than 1.61 USD and outcomes |
| Used financial services | Used to assess statistical relationship between households that used financial services in the last 12 months and outcomes |
| Access to a plot of land | Used to assess statistical relationship between households that have access to a plot of land for crops and outcomes |
| Household raises livestock/crops with intent to sell | Used to assess statistical relationship between households that raise livestock and/or crops with the intention to sell and outcomes |
| **Livestock/Crops** |  |
| Household raises oxen | Used to assess statistical relationship between households that raised oxen and outcomes |
| Household raises poultry | Used to assess statistical relationship between households that raised poultry and outcomes |
| Household raises goats | Used to assess statistical relationship between households that raised goats and outcomes |
| Household planted crops it made decisions over | Used to assess statistical relationship between households that planted crops it made decisions over and outcomes |
| **WASH** |  |
| Household has handwashing materials | Used to assess statistical relationship between households that has handwashing materials and outcomes |
| Household correctly treats water | Used to assess statistical relationship between that correctly treat water with point of use methods and outcomes |
| Household uses improved sanitation facilities | Used to assess statistical relationship between households that uses improved sanitation facilities and outcome |

